# FERROKINETIC EVALUATION OF PATIENTS WITH ACUTE CORONARY SYNDROME AT A SOUTH AMERICAN HOSPITAL, PILOT STUDY

**DOI:** 10.1101/2023.03.27.23287818

**Authors:** Luis Andres Dulcey Sarmiento, Juan Sebastián Theran Leon, Valentina Cabrera Peña, Rafael Guillermo Parales Strauch, Raimondo Caltagirone, Edgar Camilo Blanco Pimiento, María Paula Ciliberti Artavia, Juan Camilo Martinez

## Abstract

**Introduction:** Alterations in the ferrokinetic profile are associated with cardiovascular disease, and the short- and long-term prognosis of these alterations is currently unknown.

**Design:** Prospective cohort analytical observational study

**Methods:** To determine the prognostic value of the alterations of the ferrokinetic profile in patients with myocardial infarction with and without ST-segment elevation, in a Health institution from July 2017 to May 2018. Results : From **72** patients, the main affected gender was male, in ages over 56 years old, the infarction with ST elevation being the most frequent. Among the associated comorbidities, the main one was hypertension with (n: 22; 53.7%) for infarction with elevation and (n: 23; 74.2%) for infarction without ST elevation. The most frequent alteration of the parameters of the ferrokinetic profile studied, was the iron deficit, found in (n:15; 36.6%) of the patients with ST elevation and (n:13; 41.9%) without ST elevation. Low levels of hemoglobin were present at admission (n:10; 24.4%) of the subgroup with ST elevation and (n:10; 32.3%) for no ST elevation, increasing the percentage to (n:13; 31.7%) (RR: 2) (95% CI -0.131-30.63), associated with low hemoglobin values at day 7 of hospitalization. There were 2 deaths (2.77%), which presented low levels of iron without anemia and infarction with ST elevation complicated by cardiogenic shock.

**Conclusions:** iron deficiency is a common comorbidity with a high mortality rate, and the decrease in hemoglobin after hospital admission was related to mortality, so both parameters should be taken into account in Infarction and cardiovascular disease.

## Introduction

Iron is an essential element for life, since it participates in practically all oxidation-reduction processes. We can find it forming an essential part of the enzymes of the Krebs cycle, in cellular respiration and as an electron carrier in cytochromes (1). The maintenance of normal iron metabolism is particularly important for cells that are characterized by a high mitogenic potential and high energy demand (2-3), and its deficiency may be an important comorbidity in high-risk patients (4). In angiographic studies, ferritin has been associated with coronary atherosclerosis in certain populations, such as Iran (5), while it has not been in Europeans or Americans (6). Iron deficiency is the most frequent nutritional disorder of all, affecting a third of the world population (7). In other studies, anemia has been shown to be an important independent determinant of cardiovascular adverse events and death (8). In the world, more than 17 million people die each year from cardiovascular (CV) diseases (9). In our region, there are few studies that relate alterations in iron and hemoglobin values with cardiovascular (CV) diseases such as acute coronary syndrome (ACS) and their prognostic implications, so it was considered important to carry out a prospective analytical observational study., where the alterations of the ferrokinetic profile and their prognostic value in patients with ischemic heart disease in a Coronary Care Unit of a fourth level institution were determined from 07-01-17 to 05-31-18. The study will help to develop therapeutic and preventive behaviors that are not yet used in this type of patients and will contribute to contrast data with other studies from different countries.

### Ethical aspects

The realization of the present work, was adapted to the recommendations for biomedical research of the Declaration of Helsinki of the World Medical Association in its 64th General Assembly, in Fortaleza, Brazil, in October 2013 and what is contemplated in the Code of Medical Ethics of the Venezuelan Medical Federation of March 20, 1985, in its Title V, Chapter 4, referring to research in human beings.

Additionally, all patients who met the inclusion criteria and had the signed authorization to participate in the research, were informed of the purposes of the research by the research group verbally and once the family member or the patient If they were adequately informed and agreed to take part in the study, they signed the informed consent specifically designed for this research. The research was reviewed and approved by the ethics committee of the Hospital Universitario de los Andes and the Internal Medicine Specialization program.

## Materials and methods

Prospective analytical observational study.

### Aim

To determine the prognostic value of the alteration of the iron kinetic profile in patients with acute coronary syndrome (ACS): STEMI and STEMI, confined in the Coronary Unit of a IV level institution.

During the period from July 1, 2017 to May 31, 2018, all those patients with diagnoses of Acute Coronary Syndrome (ACS): Acute myocardial infarction with and without ST-segment elevation (NSTEMI and STEMI) of both genders who were admitted to the hospital were selected. the Coronary Care Unit of a level IV hospital.

The inclusion criteria were the following:

I. Patients of both genders over 18 years of age with already established clinical and paraclinical criteria for acute coronary syndrome (ACS).

Exclusion criteria:

I. Ischemic heart disease
II. Stage IV, V Chronic Kidney Disease
III. BMI > 39.9
IV. Congestive heart failure (CHF) NYHA functional class IV
V. Patient in Killip IV risk scale
VI. Diagnosed malignant neoplasm.
VII. Those who have received in the last 2 months transfusion of blood products or supplements with iron salts.
VIII. Infectious processes in any form of presentation.
IX. hemorrhagic manifestations.

Hemoglobin values were determined by the microhematocrit method, taking a blood sample from the fingertip with a capillary on admission and 7 days after hospitalization. The iron kinetic profile was determined by direct colorimetry method with Wiener lab and Bioline reagents, processed in a STAT FAX, STAT FAX MILENIUM III reader with ELISA technique and were performed on admission. The data provided by the commercial house of the test system used (table 1) were taken as normal reference values of the ferrokinetic profile.

**Table 1.** Reference values of parameters of the ferrokinetic profile

| **Table 1.** Reference values of parameters of the ferrokinetic profile |  |
| --- | --- |
| **ferrokinetic pattern** | **Normal value in adults** |
| serum iron | 49 – 181 µ/dl |
| Ferritin | 28 – 397 ng/dl |
| transferrin | 250 – 400mg/dl |
| Transferrin saturation | 20 – 55% |
| TIBC | 250 – 400 µg/dl |

### Source

Prospectus of the test system for the determination of the ferrokinetic profile of the commercial brand; Wiener lab ® and Bioline ®

The data obtained through the collection form, were processed in the SPSS version 21 program (IBM Corporation, New York, US) for Windows through which a descriptive analysis of the data was carried out, applying proportions, reasons, measures of central tendency (mean, median, mode) and measures of dispersion (range, variance, standard deviation).

The statistical association analysis was performed with the Chi-square test (□ ^2^) and the Student’s t-test, the results of which are presented in tables and graphs. The analysis to estimate the strength of association was performed by determining the relative risk. Survival analysis was performed using the Kaplan-Meier method to determine the prognostic value of changes in the ferrokinetic profile, which were presented in graphs. Statistical significance was considered for p values < 0.05. It was not possible to control the loss of subjects.

## Results

The sample obtained after meeting the inclusion and exclusion criteria was made up of 72 patients who presented acute coronary syndrome (ACS) with ages ranging from 27 to 94 years, classified into 2 subsets according to clinical and paraclinical criteria, ST-elevation infarction (STEMI) with 56.90% of the sample and 43.10% for patients with non-ST elevation acute myocardial infarction (NSTEMI) (Figure 1).

**Figure 1.**
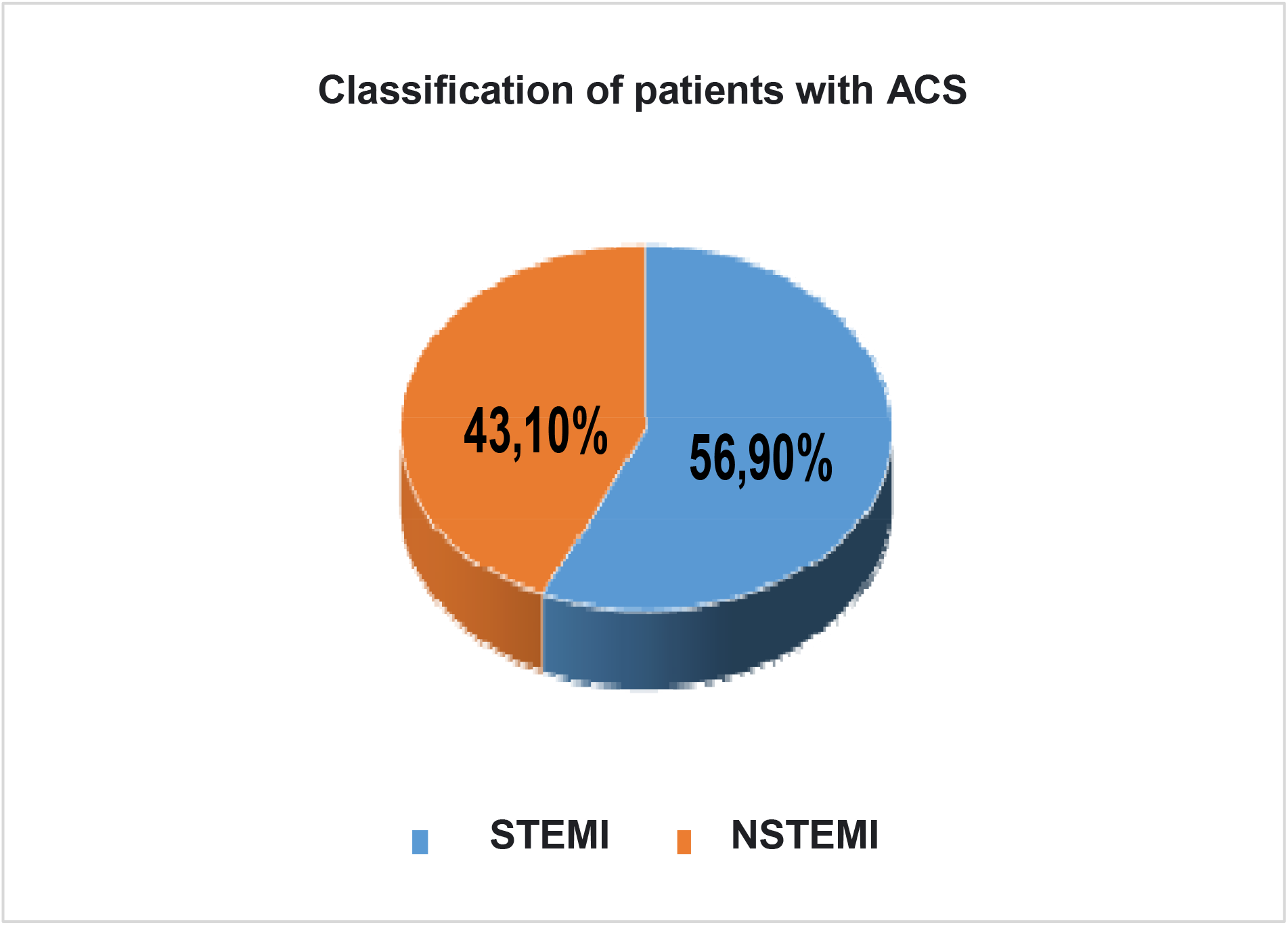
Classification of patients with ACS admitted. June 2017-May 2018

A frequency of STEMI (61%) and STEMI (54.8%) was observed in the male gender, followed by the female gender with percentages of 39% and 45.2% respectively.

## Discussion

The discussion of the results found is presented here, highlighting the important or statistically significant ones.

Table 1 showed a higher frequency of STEMI (61%) and STEMI (54.8%) in the male gender and for the female gender with percentages of 39% and 45.2% respectively.

Table 2 shows that in terms of demographic characteristics, patients between 56 and 65 years old were the ones who made up the majority of the study sample.

**Table 2.**
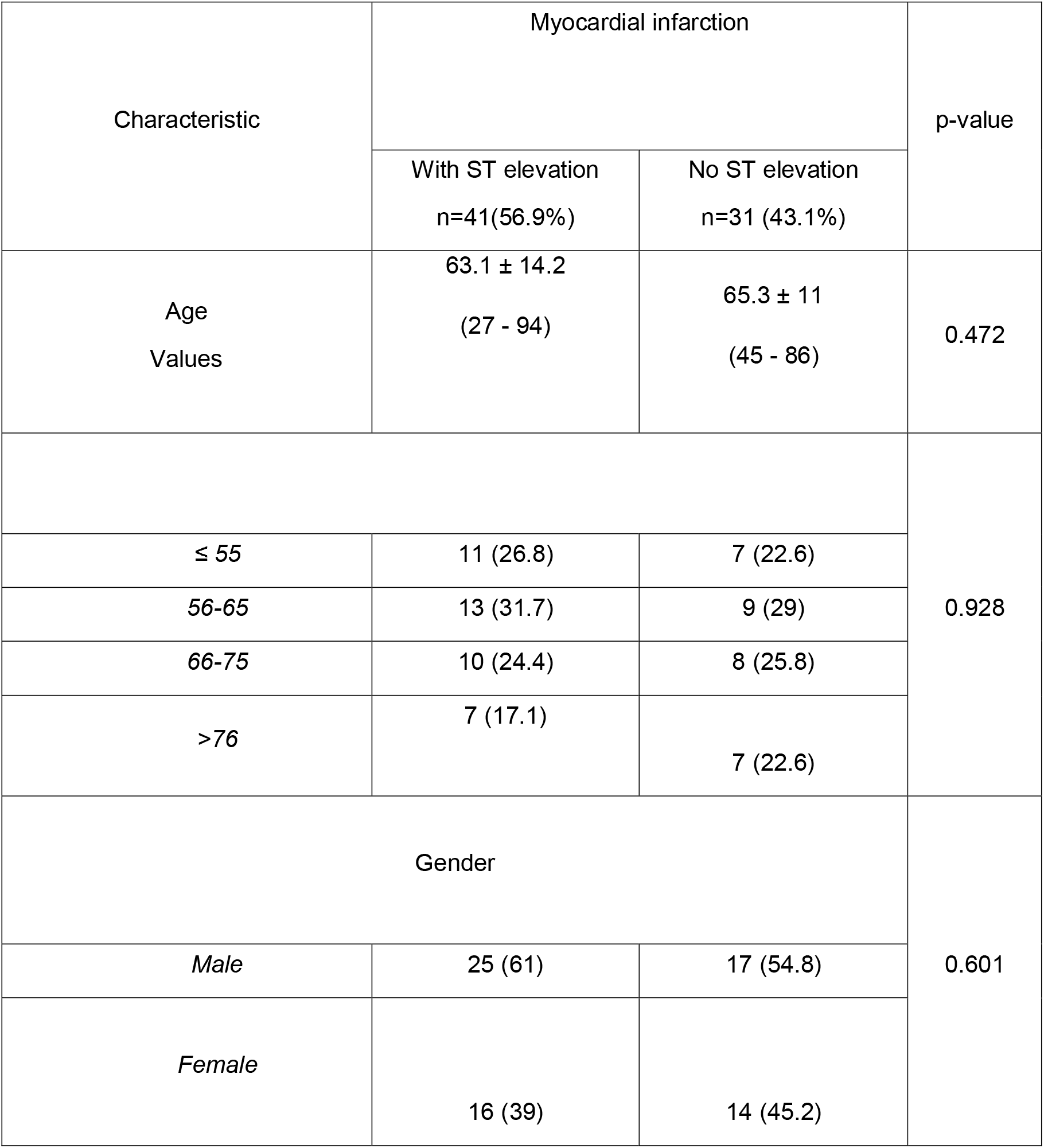
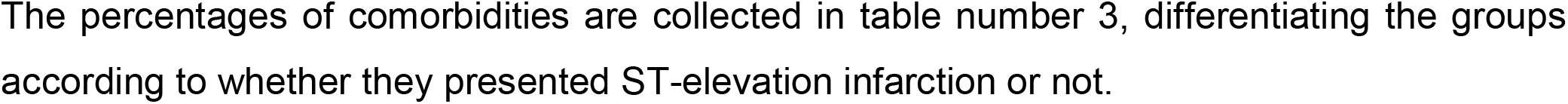
Demographic characteristics of patients with ACS admitted to the coronary care unit of a level IV institution, June 2017-May 2018.

Table 3 shows the most common comorbidities in these patients, the main one being arterial hypertension with a frequency of 53.7% for patients with STEMI and 74.2% in patients with STEMI.

**Table 3.**
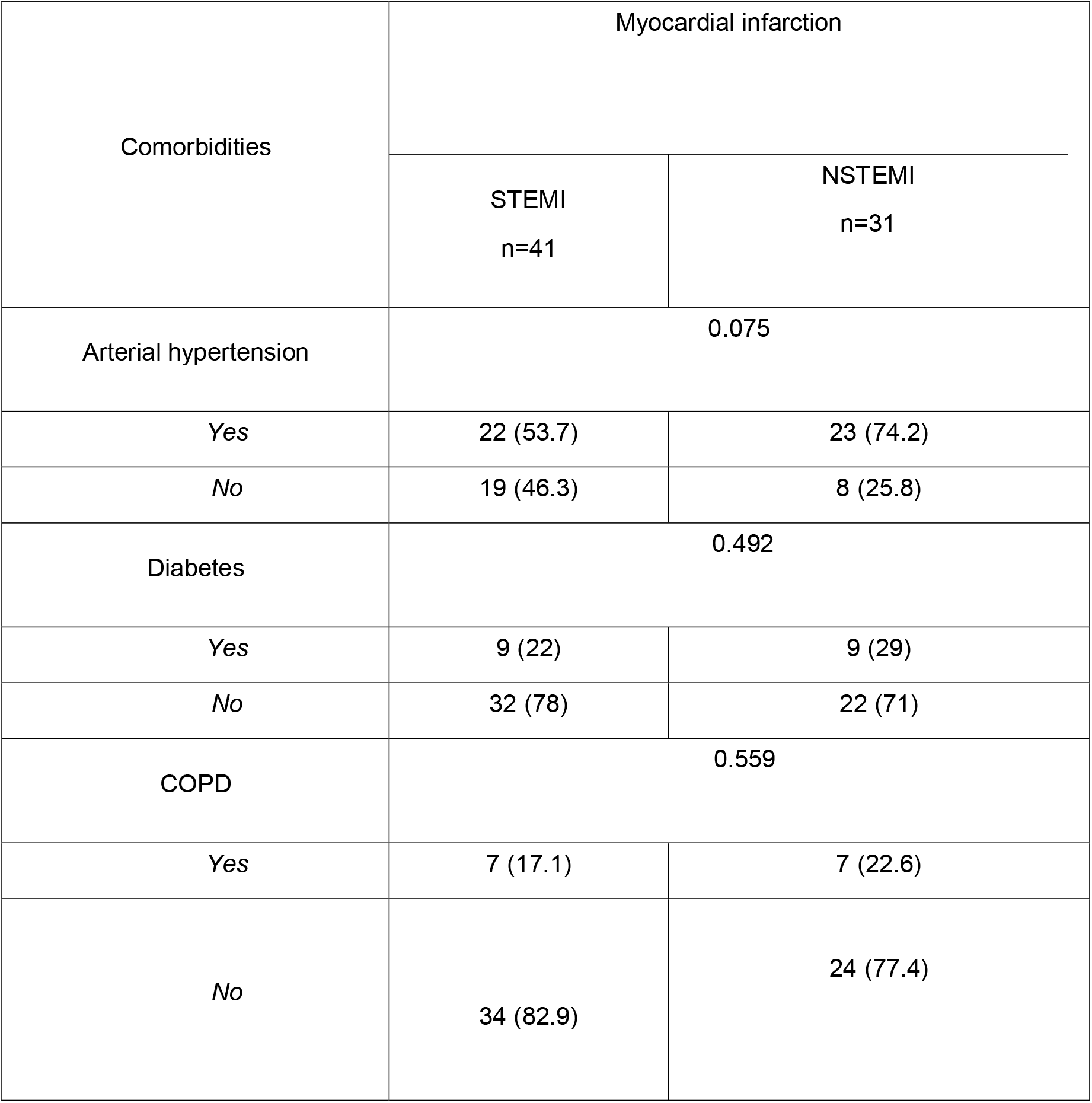
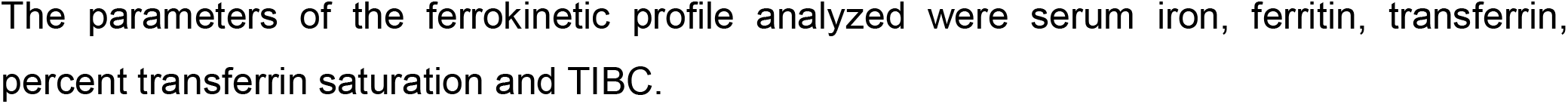
Comorbidities of patients with ACS admitted to the coronary care unit of a level IV hospital, June 2017-May 2018.

In Table 4, regarding the parameters of the ferrokinetic profile analyzed, in patients with STEMI it was evidenced that iron deficiency was the most frequent ferrokinetic alteration at 36.6%, followed by alterations in transferrin and saturation percentage. of transferrin with 22% and ferritin 17.1%. Additionally, it was found that in patients with NSTEMI, iron deficiency was the main disorder in 41.9% of the patients in the sample, followed by the percentage of transferrin saturation for 22.6%. Hemoglobin values at hospital admission were low in 24.4% of the patients with STEMI and in 32.3% with STEMI. Hemoglobin levels decreased on the seventh day of hospital stay, by 31.7% for patients with STEMI.

**Table 4.**
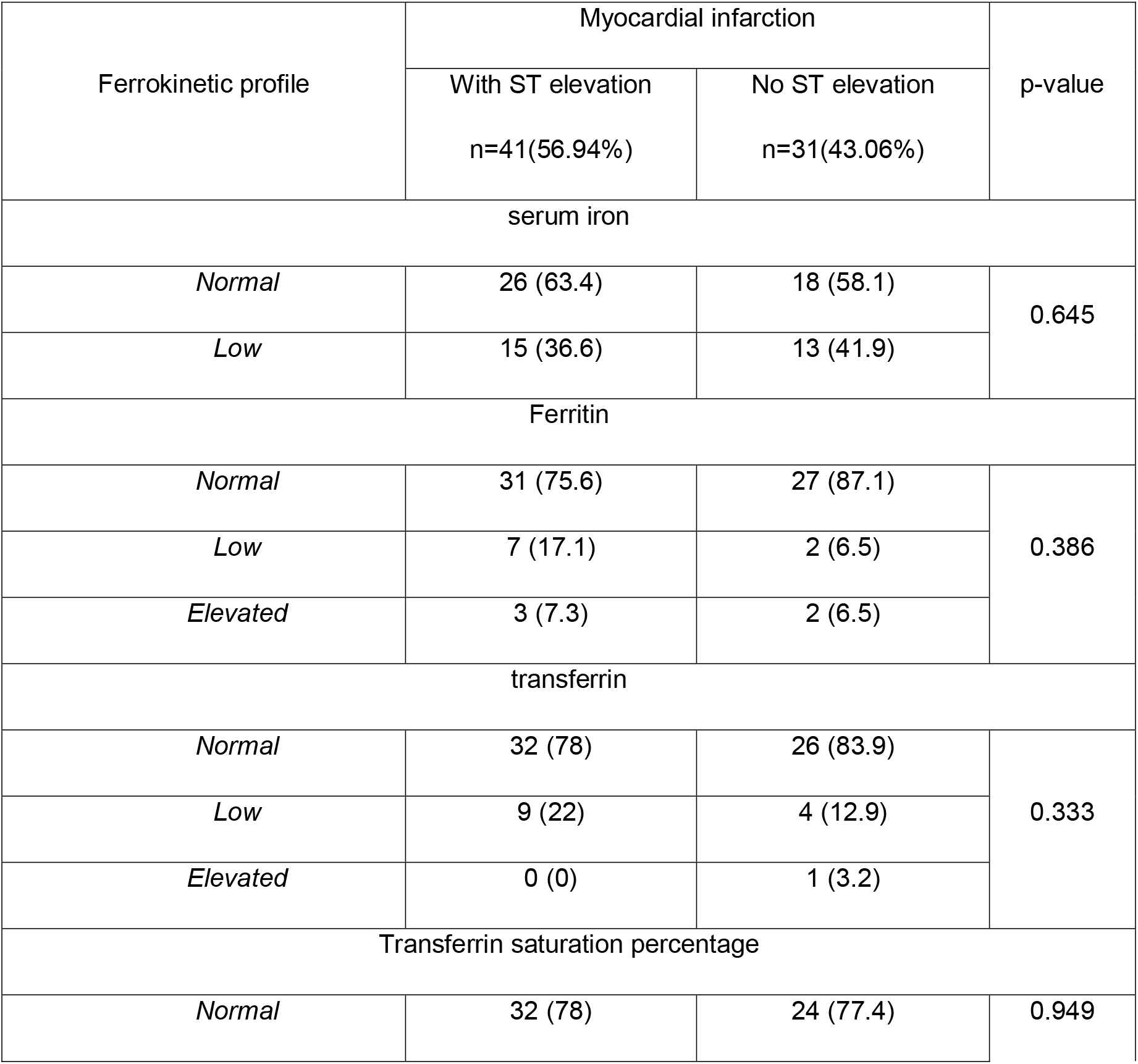
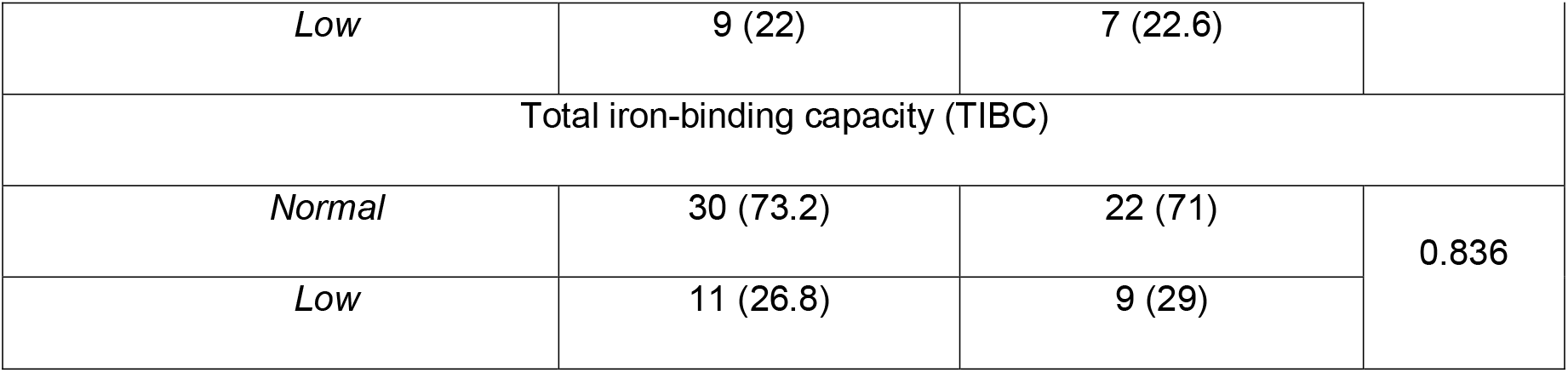
Ferrokinetic profile of patients with ACS admitted to the coronary care unit of a level IV hospital June 2017-May 2018.

In Table 5, when evaluating the hemoglobin values at admission, a higher frequency of anemia was found in the male gender with 65% of the total patients and 35% for the female gender, the frequency being higher in the group age between 56 - 65 years.

**Table 5.**
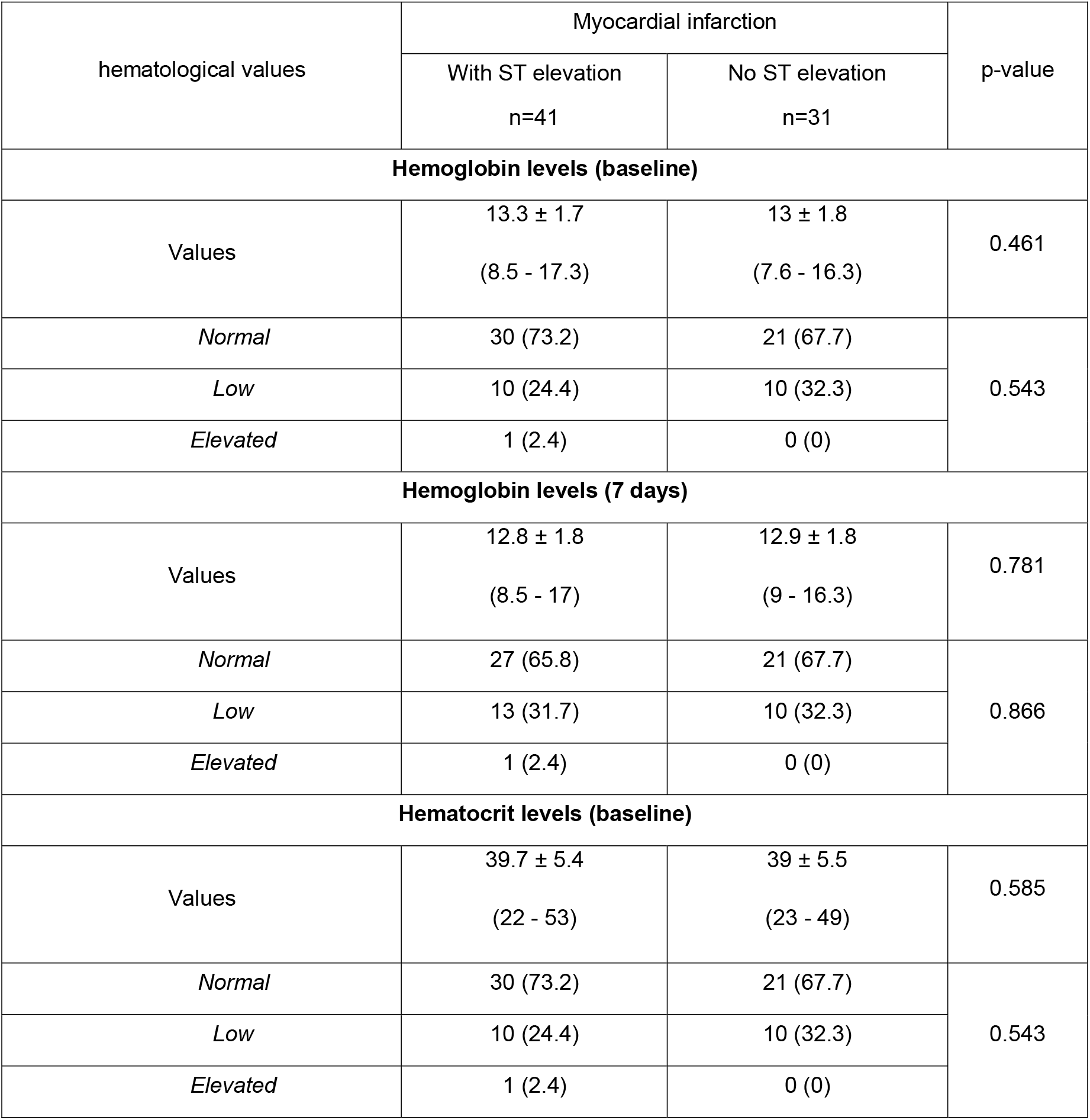
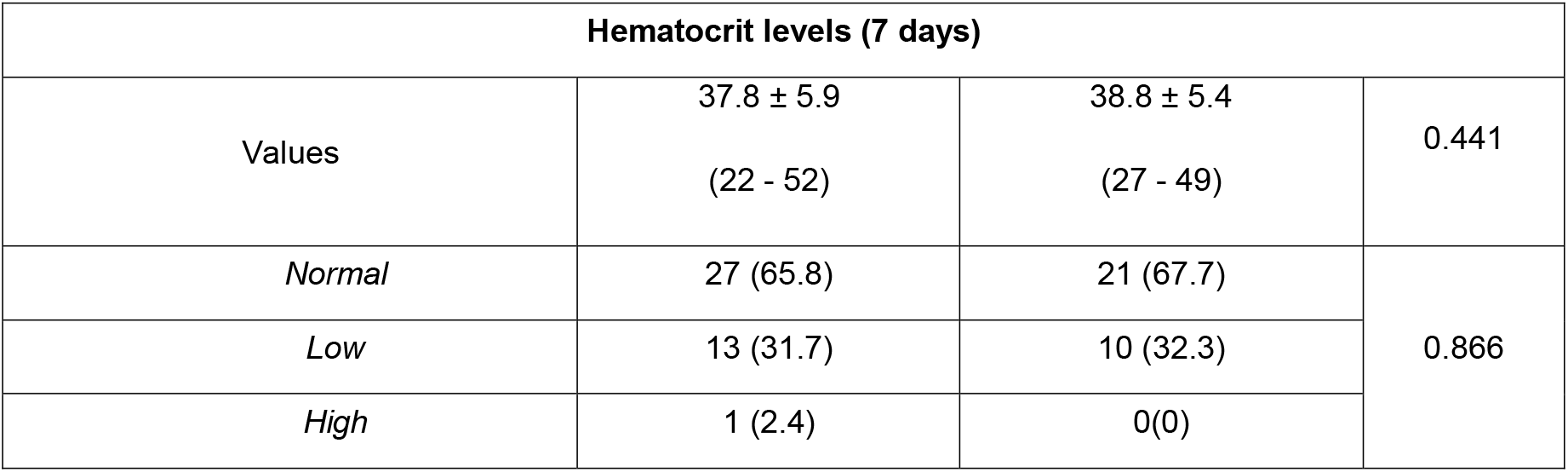
Hemoglobin and hematocrit values at admission and on day 7 of patients with ACS admitted to a level IV hospital June 2017-May 2018.

Regarding the classification of the left ventricular ejection fraction (LVEF) according to the 2016 heart failure guidelines of the European Society of Cardiology (SEC), it was found to be preserved in both subsets, with 56.1%, n□ 23, in patients with STEMI and 64.5%, n□ 20 in STEMI (Figure 2).

**Figure 2.**
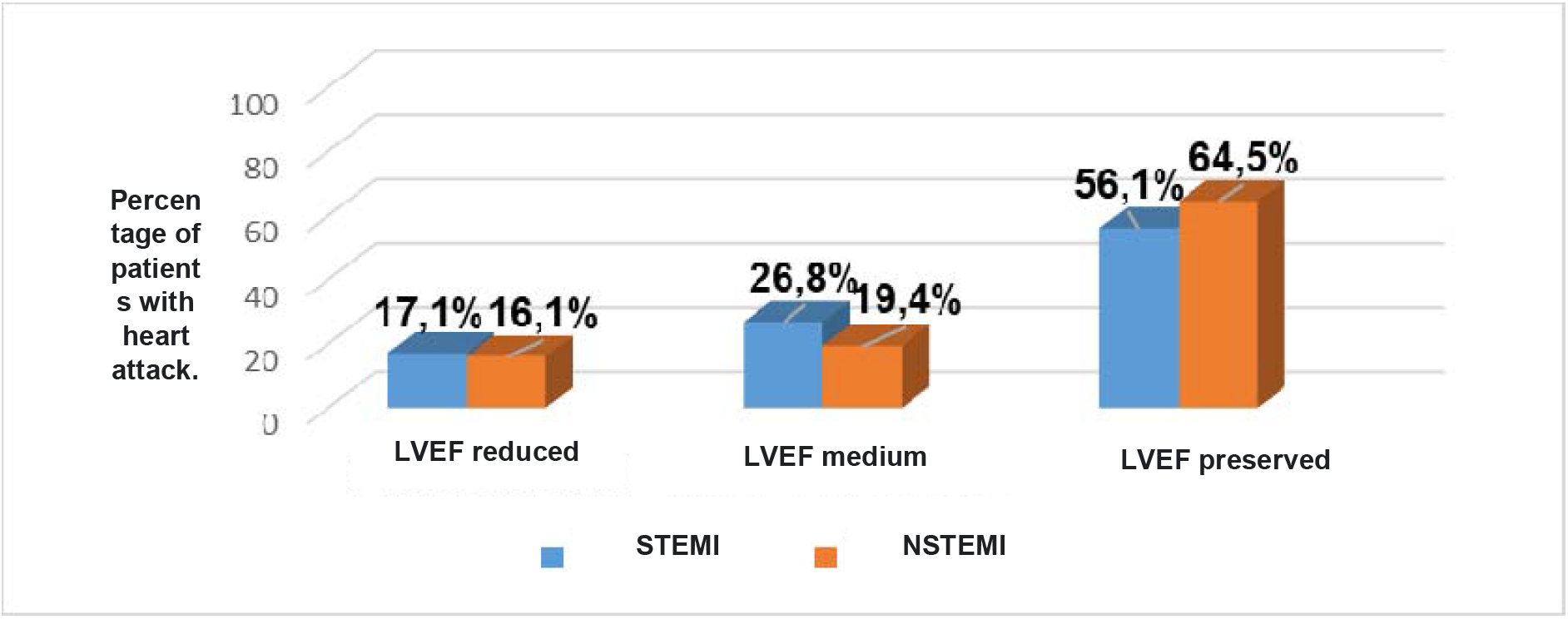
Classification of the left ventricular ejection fraction (LVEF) according to the SEC in patients with ACS admitted to the coronary unit of a IV level hospital, June 2017-May 2018.

In Table 6, LVEF correlated with serum iron levels was found preserved in 65.9% of patients with normal iron levels, LVEF in the intermediate range in 22.7% in those with normal iron, LVEF was found reduced in 11.4% in those with low iron respectively, not showing statistical significance.

**Table 6.**
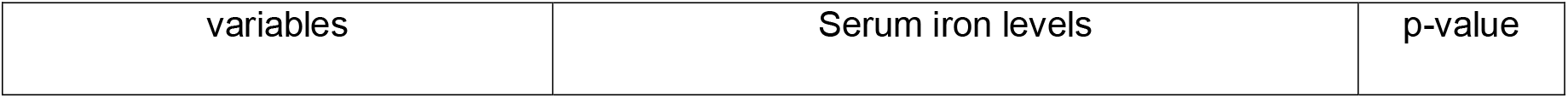
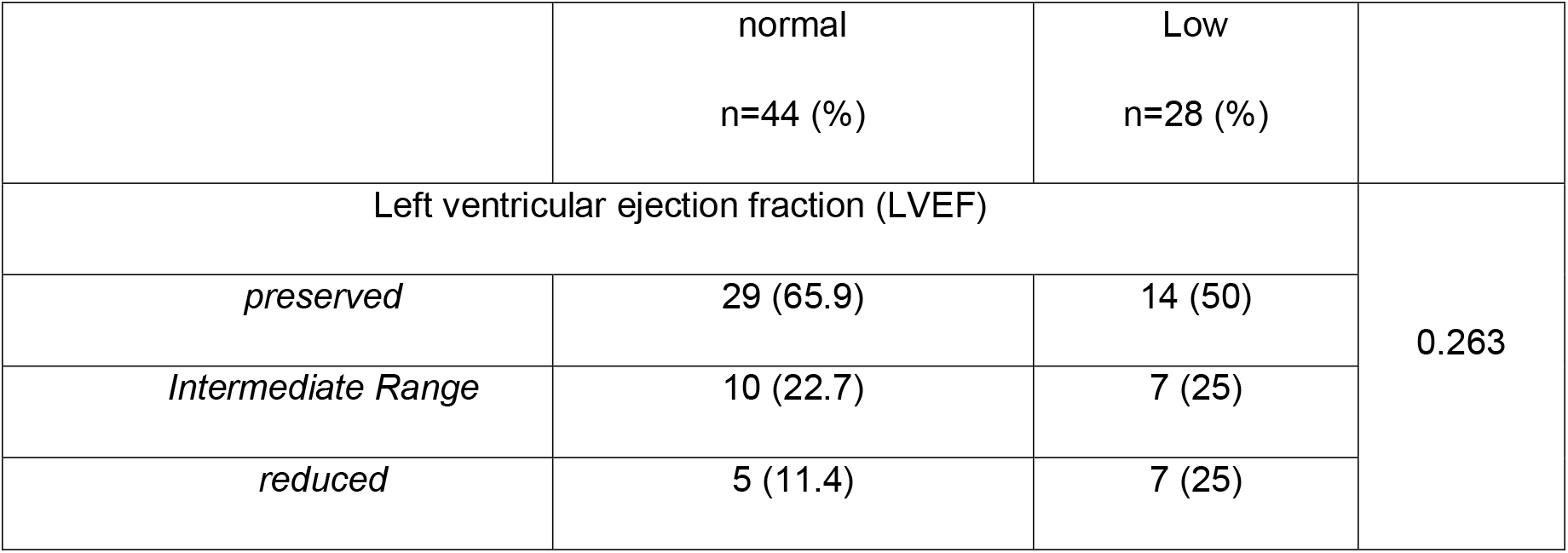
Correlation of serum iron levels with LVEF in patients with ACS admitted to the coronary unit of the IV level hospital from June 2017 to May 2018.

In Table 7, when correlating LVEF with ferritin, it was observed that LVEF was preserved in 70% of those with normal ferritin; in the group with low ferritin, LVEF was in the intermediate range, and LVEF was reduced in 40% of patients with excess of this ferrokinetic parameter with a p value □ 0.011.

**Table 7.** Correlation of ferritin levels with LVEF in patients with ACS admitted to the coronary unit of the IV level hospital, June 2017-May 2018.

| Values | ferritin levels |  |  | p-value |
| --- | --- | --- | --- | --- |
|  | normal<br>n=58 (80.5%) | Low<br>n=9 (12.6%) | elevated<br>n=5 (6.9%) |  |
| Left ventricular ejection fraction |  |  |  | 0.011 |
| *preserved* | 40 (70) | 1 (10) | 2 (40) |  |
| *Intermediate* | 11 (20) | 5 (60) | 1 (20) |  |
| *reduced* | 7 (10) | 3 (30) | 2 (40) |  |

In Table 8, when the correlation between low iron levels and mortality was made, a relative risk of 7.758 (95%CI 0.387-181.77) was obtained for mortality with low iron and of 0.907 (95%CI 0.59-1.385). for STEMI in the group of patients with normal iron.

**Table 8.**
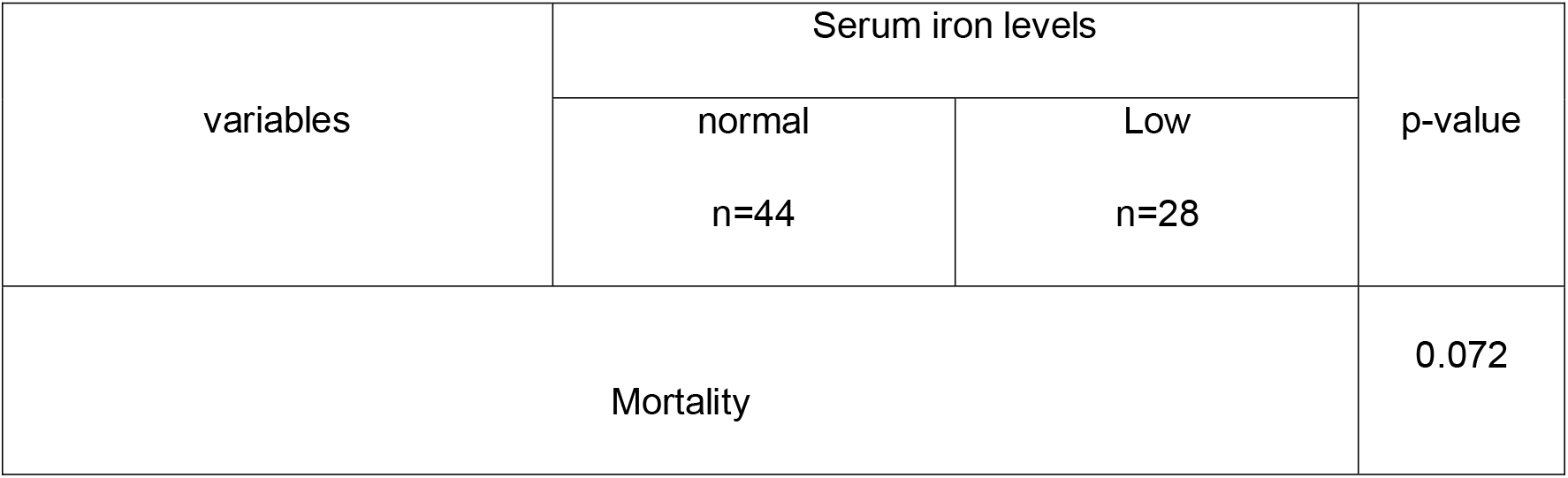
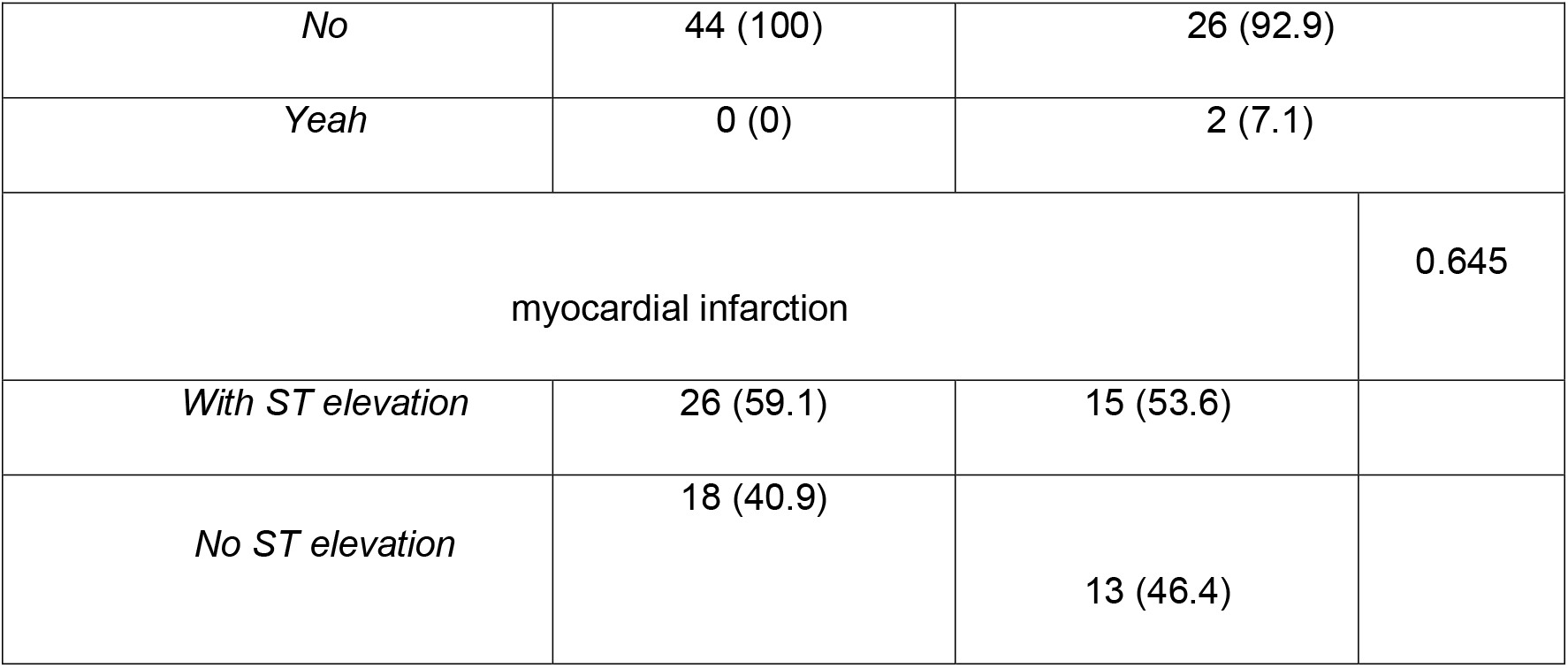
Correlation of serum iron levels with hospital mortality in patients with ACS admitted to the unit of a level IV hospital June 2017-May 2018.

A relative risk of 0.504 (95% CI 0.022-10.772) was observed for mortality in the group of patients with low hemoglobin at admission and a relative risk of 2 (95% CI 0.131-30.630) for mortality in the group of patients with low hemoglobin at admission. day 7 of hospitalization.

In Figure 3, survival prognosis was evaluated using the Kaplan-Meier statistical method, during a 7-day follow-up, after hospital admission, using a survival curve, whose differences were correlated with the Log-Rank test (Mantel - Cox), determining a statistical value of p =0.157. The indirect risk of death (odds ratio) was estimated at 8.4 (95% CI: 0.4 – 181.8) for low iron (p = 0.176). Regarding the limitations found, the limited number of the sample stands out, which can be evidenced, for example, in the width of the values in the confidence intervals.

**Figure 3.**
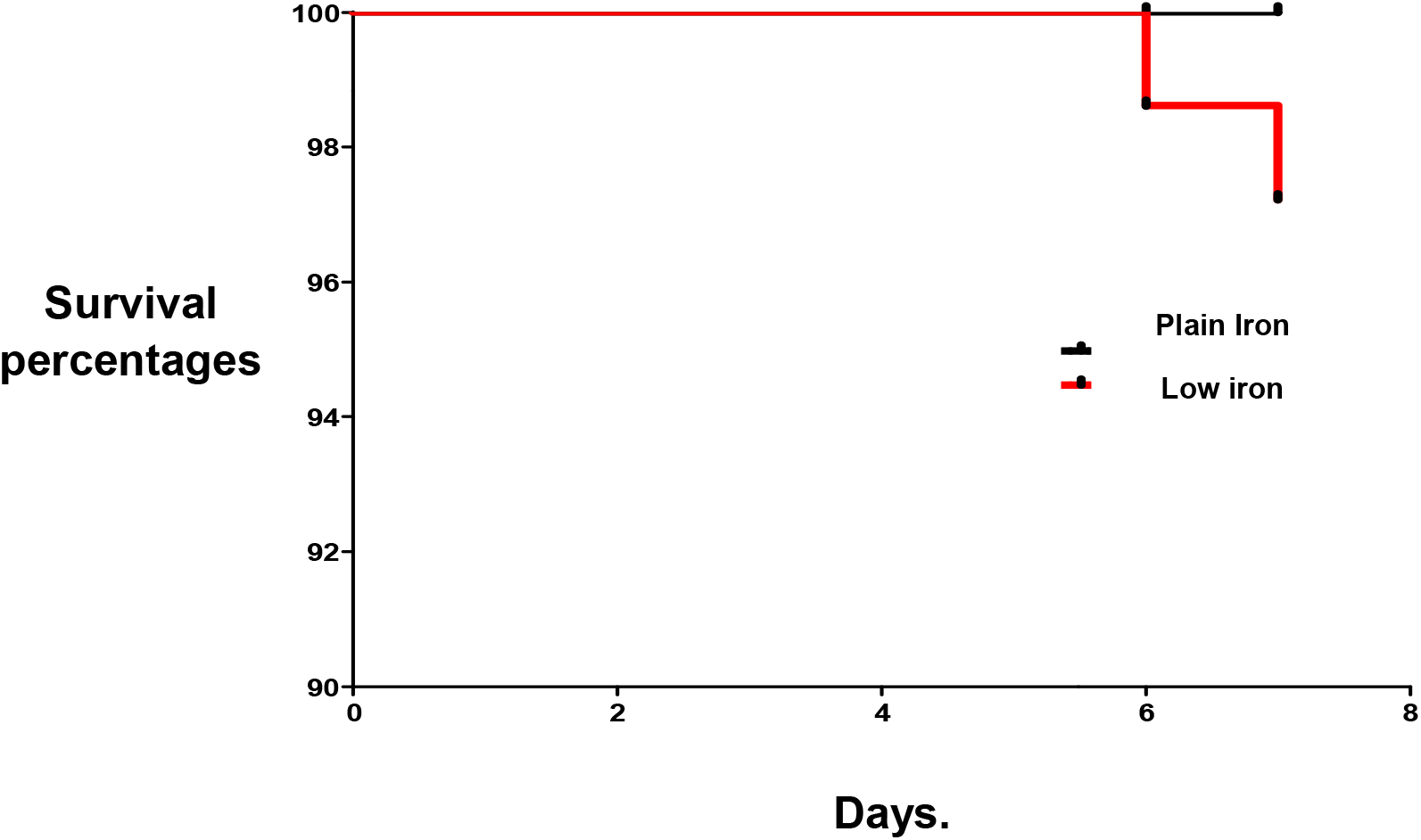
Distribution of survival (Kaplan - Meier) of patients with ACS and iron deficiency admitted to the coronary unit of a IV level hospital from June 2017 to May 2018.

It should be noted that multivariate analysis is the set of statistical methods whose purpose is to simultaneously analyze multiple data sets in the sense that there are several variables measured for each individual or object studied. Finally, this type of analysis just the differences between a large number of factors related to the result through mathematical models.

This type of multivariable analysis is suitable for adjusting for many measured confounding factors and estimates the effects of all measured confounding factors on the result, however it requires a large sample size if there are a large number of variables, since our study had a sample of 72 patients for a highly prevalent disease, which creates a context of bias that makes it necessary to carry out studies with larger populations.

Similarly, in our study, the absence of a statistical analysis to control confounding and interaction variables here means that the results and conclusions presented could have been affected.

## Conclusions

When representing the data from this study, it was found that the main gender with ACS was male, in ages between 56-65 years, which agrees with what was demonstrated by Meroño et al (10), where said clinical condition was present mainly in the masculine gender. Among the comorbidities associated with ACS in the different subsets, the main one was arterial hypertension with 53.7% for STEMI and 74.2% in STEMI, similar to what was achieved by Ponikowska et al (11).

The most frequent alteration of the parameters of the ferrokinetic profile studied was iron deficiency, found in 36.6% of the patients with STEMI and 41.9% of the STEMI, this clinical condition being prevalent in these patients as verified in the study by González et al (12) also in patients with ACS, likewise low ferritin levels were the second most frequent alteration in this study, as reported by Archbold et al (13) in their article, both serum ferritin and transferrin predicted an association of all-cause mortality.

Low hemoglobin levels were present at hospital admission in 24.42% of the subgroup with STEMI and 32.30% for STEMI, increasing the percentage to 31.7% in the first subgroup with a relative risk (RR) of 2 (CI95 % 0.131-30.63), regarding the decrease in hemoglobin on day 7 of hospitalization, this situation was independently associated with a higher incidence of adverse events, as was also demonstrated by Carberry et al (14) and Colombo et al (15).

The male gender presented the highest frequency of low hemoglobin values at hospital admission with 65% and 35% for the female gender, differing this finding with what is found in the current literature, where the highest prevalence was attributed to the female gender in that reported in the study by Carberry et al (14), with anemia being a powerful predictor of major adverse cardiovascular events in these patients similar to that observed in the study by Acharya et al (16).

In our study, there were no bleeding complications that justified anemia upon admission or during the hospital stay, possibly associated with blood extractions, inadvertent digestive bleeding, hematopoietic deficiency disorders, or other disorders that would undoubtedly contribute to anemia (17).

When the evaluation of low iron levels by gender and age group is carried out, it is evident that the female gender presented the highest frequency of this condition in 57% with a statistical significance of p□ value 0.034, equal to what was demonstrated in other series of reviews, where it is presented as a frequent comorbidity of cardiovascular diseases (18), with ages between 66 and 75 years being the most prevalent with 50% of the sample, with statistical significance with p value □ 0.0001, similar to the sociodemographic results obtained in another study (19).

When the correlation of comorbidities with iron deficiency was carried out, it was observed that hypertension was present in 71.40% of patients with low serum iron, followed by COPD in 32.10% of said group, with a statistically significant value. (p□0.030), this finding in our study when correlated with that found by Hsu et al (19) was very similar, where iron deficiency presented a statistically significant result for the COPD variable (p 0.010), however in the In the case of the present study (19), hypertension and its association with the iron deficiency variable showed a statistical value of (p < 0.001).

The LVEF correlated with the serum iron levels could be appreciated, it was found preserved in 65.9% of the patients with normal iron levels, the LVEF in the intermediate range in 22.7% in those with normal iron and the LVEF was found reduced in 11.4 % in those with low iron, respectively, not showing statistical significance. These results are not similar to what was demonstrated in the study by Huang (20) where iron deficiency was associated with a reduction in Functional Capacity and a greater risk of adverse events. in patients with heart failure.

Regarding the correlation of LVEF with ferritin, LVEF was preserved in 70% of those with normal ferritin, in the group with low ferritin, LVEF was in the intermediate range, and LVEF was reduced in 40% of patients. with an excess of this ferrokinetic parameter with a p value □ 0.011, these results contrast with what was reported by Weidmann (21) where ferritin concentrations were not significant in acute myocardial infarction and showed no relationship with ventricular ejection fraction either. left post infarction.

In the first 7 days of hospitalization, 2 fatalities occurred, representing 2.77% of the total sample, which presented low iron levels without anemia and STEMI, these deceased presented complications such as complete AV block and cardiogenic shock, contrasting with what has been described. in the literature where mortality in myocardial infarction with ferrokinetic alterations was correlated with the presence of anemia (14). In our study, mortality occurred in 7.1% of the patients with iron deficiency, obtaining a relative risk of 7.758 (95% CI 0.387-181.77) of mortality for the subgroup with STEMI, this result being a similar important determinant. to what was observed in other studies (22) where alterations in iron levels strongly predicted cardiovascular disease and all-cause mortality independent of other variables, this being possibly associated with high levels of inflammatory mediators such as ferritin or other factors previously mentioned. (23-25).

When evaluating the prognosis during the first 7 days, associated with changes in the iron kinetic profile, the main disorder found was iron deficiency, determining survival using the Kaplan-Meier survival method, the difference between the curves was evaluated with the Log-Rank test (Mantel-Cox), finding a value of p = 0.157. The indirect risk of death (odds ratio) was estimated at 8.4 (95% CI: 0.4 – 181.8) for low iron (p = 0.176). Given the non-availability of studies, reviews or meta-analyses where the prognostic value of iron in the acute phase of ACS in Latin America is evaluated, there are no doubts about this topic to be investigated in larger population groups with sufficient statistical power to answer this question..

In patients with ACS, iron deficiency is a very frequent comorbidity with a high mortality ratio, likewise the decrease in hemoglobin values after hospital admission correlates significantly with mortality, so both parameters in the future after being validated could be taken into consideration as determinants of prognostic stratification in the group of patients with acute or chronic cardiovascular disease. We are self-critical with the results reported here, since the biases mentioned regarding the limited number of the sample that reflects the wideness of the values of the confidence intervals, the lack of a multivariate study and the absence of a statistical analysis for control of confounding variables and interaction with necessary in order to generalize and validate what is reported here. With this study, a call is made to develop this type of research in other latitudes that contribute to understanding factors that up to now have not been taken into account in the prognosis of cardiovascular diseases, both in their acute and chronic phases.

## Data Availability

All data produced in the present study are available upon reasonable request to the authors

